# Exercise and brain health in patients with coronary artery disease: study protocol for the HEART-BRAIN randomized controlled trial

**DOI:** 10.1101/2024.05.22.24307744

**Authors:** Angel Toval, Patricio Solis-Urra, Esmée A Bakker, Lucía Sánchez-Aranda, Javier Fernández-Ortega, Carlos Prieto, Rosa María Alonso-Cuenca, Alberto González-García, Isabel Martín-Fuentes, Beatriz Fernandez-Gamez, Marcos Olvera-Rojas, Andrea Coca- Pulido, Darío Bellón, Alessandro Sclafani, Javier Sanchez-Martinez, Ricardo Rivera-López, Norberto Herrera-Gómez, Rafael Peñafiel-Burkhardt, Víctor López-Espinosa, Sara Corpas- Pérez, María Belén García-Ortega, Alejandro Vega-Cordoba, Emilio J. Barranco-Moreno, Francisco J. Morales-Navarro, Raúl Nieves, Alfredo Caro-Rus, Francisco J. Amaro-Gahete, Jose Mora-Gonzalez, Sol Vidal-Almela, Anna Carlén, Jairo H. Migueles, Kirk I. Erickson, Eduardo Moreno-Escobar, Rocío García-Orta, Irene Esteban-Cornejo, Francisco B. Ortega

**Author notes:** Correspondence: Corresponding authors: Francisco B. Ortega and Angel Toval. Department of Physical Education and Sports, Faculty of Sports Science, University of Granada; Carretera de Alfacar, 21. Granada 18071, Spain; and. **Trial Sponsor:** University of Granada; Crtra. Alfacar, 21. Granada 18071, Spain. **Trial registration:** ClinicalTrials.gov: NCT06214624.

## Abstract

****Introduction**:** Patients with coronary artery disease (CAD), also called coronary heart disease, have a higher risk of developing cognitive impairment and mental health disorders compared to the general population. There is a need to identify effective and sustainable strategies to improve brain health in individuals with CAD, in which physical exercise could play a major role. The overall goal of the HEART-BRAIN randomized controlled trial (RCT) is to investigate the effects of exercise, including different types, on brain health outcomes in patients with CAD, and the underlying mechanisms.

****Methods**:** This three-arm, single-blinded RCT will include 90 adults with CAD, aged 50-75 years. The participants will be randomized into: 1) control group - usual care (n=30), including periodic medical visits and medication management, 2) aerobic high-intensity interval training (HIIT) (n=30), or 3) aerobic HIIT combined with resistance exercise training (n=30). The intervention will last 12 weeks, offering 3 sessions (45min each) per week to the exercise groups, and the study outcomes will be assessed at baseline and after the intervention. The primary outcome of the study is to determine changes in global and regional cerebral blood flow assessed by magnetic resonance imaging. Secondary outcomes include changes in brain vascularization, cognitive measures (i.e., general cognition, executive function and episodic memory), and cardiorespiratory fitness. Additional health-related outcomes will be evaluated, and several potential mediators and moderators will be investigated (i.e., brain structure and function, cardiovascular and brain-based biomarkers, hemodynamics, physical function, body composition, mental health, and lifestyle behavior).

****Conclusions**:** The HEART-BRAIN RCT will provide novel insights on how exercise can impact brain health in patients with CAD and the potential mechanisms explaining the heart-brain connection, such as changes in cerebral blood flow. The results might have important clinical implications by increasing the evidence on the effectiveness of exercise-based preventive strategies that could delay cognitive decline in this high-risk CAD population. Our findings will be relevant for patients with CAD, researchers and healthcare providers involved in CAD-related clinical care.

## 1 INTRODUCTION

More than 300 million people globally are currently living with coronary artery disease (CAD), also called coronary heart disease, (1), which consists of a narrowing or blockage of the coronary arteries mainly caused by atherosclerosis. CAD is the most prevalent cardiovascular disease and a leading cause of mortality and morbidity worldwide (1–5). CAD incurs a significant economic burden, with annual costs estimated at €77 billion in the EU and $239.9 billion in the USA (3, 6). Recent studies have shown that individuals with CAD are at high risk of accelerated decline in cognitive and mental health compared to aged-matched adults without CAD, supporting a nexus between the heart and the brain (7–11). Indeed, the cardiovascular risk factors associated with CAD similarly predispose individuals to cerebrovascular disease, in turn affecting their cerebral perfusion. While pharmaceutical therapies have traditionally been the cornerstone of CAD treatment, exercise is an effective non-pharmaceutical intervention for improving cardiometabolic health in these patients (12–15). However, the potential effect of exercise on brain health among individuals with CAD, as well as the mechanisms underlying the heart-brain connection, remain unknown.

A key physiological mechanism modulating the heart-brain connection might involve the intricate machinery responsible for supplying blood to the brain. Chronic cerebral hypoperfusion, marked by reduced cerebral blood flow (CBF), impedes the brain’s access to vital oxygen and nutrients (9, 16–18). Therefore, a disrupted CBF could play an important role by accelerating cognitive decline and increasing the risk of dementia and mental health disorders in patients with CAD. In turn, exercise could serve as a pivotal factor to enhance CBF, offering a promising non-pharmaceutical adjunct therapy. Limited evidence suggests that exercise may increase CBF and induce brain angiogenesis (i.e., growth of new capillaries). However, these findings are mainly derived from observational evidence and animal studies (19, 20). These hypotheses need to be tested in randomized controlled trials (RCTs) in humans. The most advanced neuroimaging techniques (particularly in magnetic resonance imaging, MRI) allow non-invasive (no contrast) methods to assess *in vivo* CBF using the Arterial Spin Labelled sequence (ASL-MRI) and cerebral vascularization using magnetic resonance angiography (MRA) (21). These state-of-the-art techniques can provide unique and novel information about the connection of these two possible mechanisms (i.e. changes in CBF and cerebral vascularization) with cognition and mental health.

Physical exercise is strongly recommended in the clinical management of patients with CAD to reduce disease progression and the risk of major cardiovascular events (13, 22). However, the mechanisms by which different types and doses of exercise induce a variety of physiological responses are still not fully understood. For instance, modalities such as high-intensity interval training (HIIT) or its combination with resistance training are understudied. HIIT may offer superior health benefits, such as larger improvements in cardiorespiratory fitness, compared to moderate intensity continuous training (MICT) in patients with CAD (8–11). However, it is unknown if HIIT is also superior for improving brain health outcomes. On the other hand, resistance training and the combination of aerobic and resistance training remain understudied, despite recommendations by the World Health Organization (WHO) (23) and the European and American clinical guidelines for CAD management (12) (14, 24), all advocating for the combination of aerobic and resistance training for optimal health benefits (12, 13). Since one of the most documented benefits of exercise is the improvement in cardiorespiratory fitness (25–27), and this marker has been consistently linked to better brain health outcomes (28, 29), cardiorespiratory fitness might serve as a potential mediator of the beneficial effects of exercise on the HEART-BRAIN outcomes.

Based on the existing research gaps, we have designed the HEART-BRAIN project, an RCT to investigate the effects of exercise training on brain health outcomes in patients with CAD, including the underlying mechanisms of the heart-brain connection. Herein, we describe the rationale and methodology of the HEART-BRAIN RCT.

### 1.1 Study objectives

The overall objective of the HEART-BRAIN trial is to investigate the effects of exercise on brain health outcomes in patients with CAD.

The <u>primary objective</u> of the study is to investigate the effects of aerobic HIIT and aerobic HIIT plus resistance training compared to usual care (i.e. no exercise training) on global and regional CBF in individuals with CAD.

<u>Secondary objectives</u>: to determine the effects of aerobic HIIT, and aerobic HIIT plus resistance training compared to usual care on (i) cerebral vascularization, (ii) executive function and general cognition, and (iii) cardiorespiratory fitness. Additionally, we will investigate the role of potential moderators and mediators (see details in statistical analysis section) on the expected effects.

<u>Tertiary objectives:</u> to determine the effects of aerobic HIIT, and aerobic HIIT plus resistance training, compared to usual care on additional outcomes related to cardiovascular and brain health, such as brain structure and function, inflammatory, cardiovascular and brain-based biomarkers, cardiac, vascular and transcranial non-invasive hemodynamics, physical function, body composition, mental health and lifestyle behavior.

Our <u>primary hypothesis</u> is that both exercise groups will have a greater increase in CBF compared to the usual care control group, in which HIIT plus resistance training might have a larger effect size than HIIT only.

## 2 METHODS AND ANALYSIS

### 2.1 Design and ethics

The HEART-BRAIN trial is a single-centered, three-armed, single-blinded RCT, in which 90 adults with CAD, between 50-75 years old, living in Granada (Spain), will be enrolled (**Figure 1)**. Participants meeting the eligibility criteria will be randomized into three groups: 1) aerobic HIIT exercise program (n=30), 2) aerobic HIIT plus resistance training exercise program (n=30), and 3) control group - usual care (n=30). The group receiving usual care will be treated as standard care in Spain which includes periodic medical revisions and medication control. The two exercise groups will undertake a 12-week supervised exercise program. All the study outcomes will be assessed at baseline and during the 12-week follow-up. Lifestyle behaviors will also be evaluated at mid-point (6 weeks).

**Figure 1.**
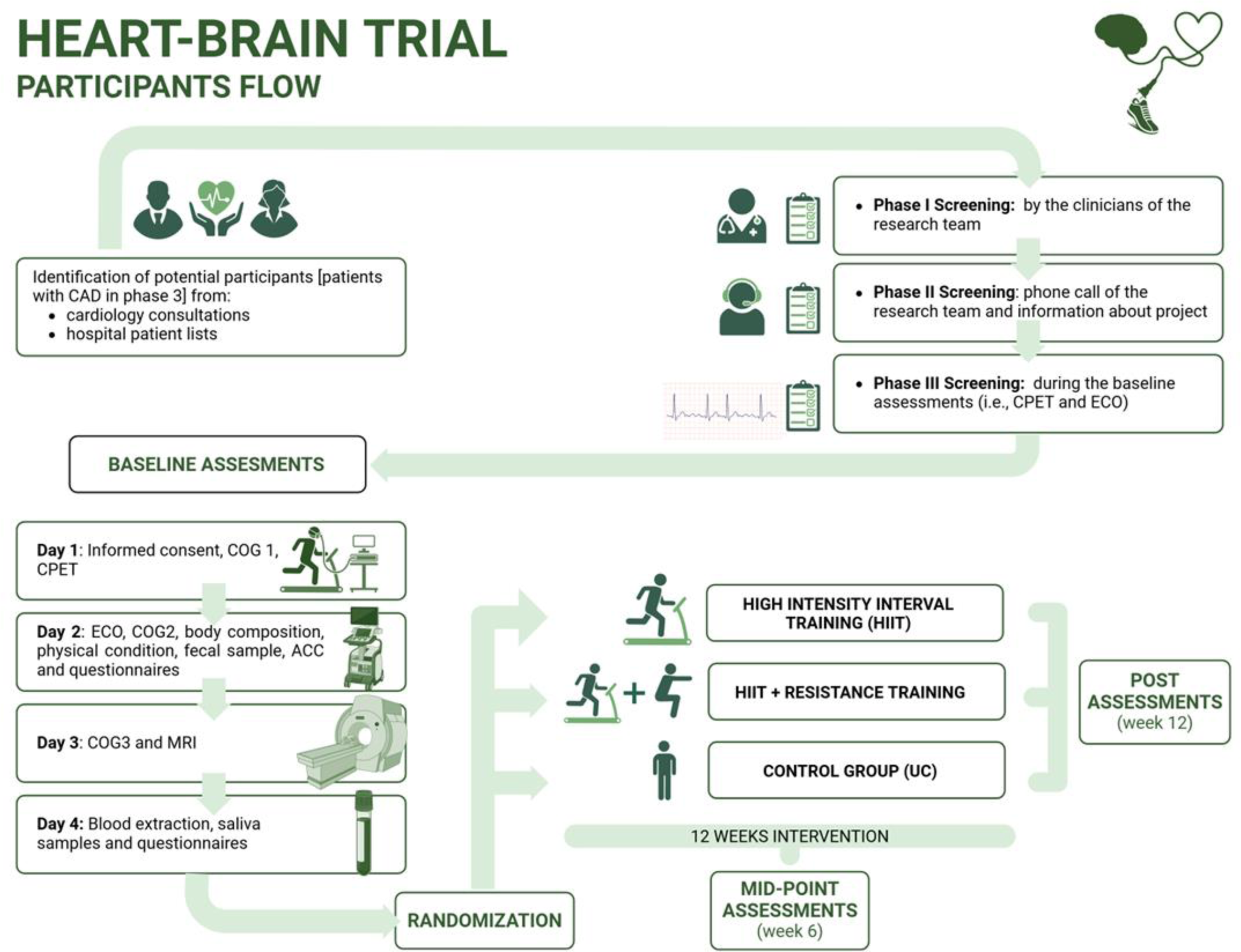
Visual representation of the study design in the HEART-BRAIN trial. ACC: Accelerometry; CAD: coronary artery disease; CPET: cardiopulmonary exercise test; ECO: echography; COG; cognitive function tests; MRI: magnetic resonance imaging UC: usual care. *Post-intervention assessment includes similar outcomes compared to baseline assessment.

Eligible participants will receive written information about the study objectives and design, and will provide informed consent before participation. The trial protocol is in accordance with the principles of the Declaration of Helsinki and was approved by the Research Ethics Board of the Andalusian Health Service (CEIM/CEI Provincial de Granada; #1776-N-21 on December 21^st^, 2021). The RCT is registered on Clinicaltrials.gov (NCT06214624; submission date: December 22^nd^, 2023) and the study protocol and statistical analysis plan were uploaded before the last participants were randomized (submission date: January 29^th^, 2024). The study has been designed following the Standard Protocol Items for Randomized Interventional Trials (SPIRIT) (30, 31) and the SPIRIT-Outcomes 2022 Extension (32) (**Supplementary Table 1**). Any significant changes to the protocol will be reported to the trial registry and the Research Ethics Board, which is in line with the SPIRIT guidelines. Moreover, participants in the HEART-BRAIN trial are insured under a social responsibility policy from the University of Granada, which covers ancillary services, post-trial care or compensation if necessary. There are no financial rewards provided to participants for their involvement in the study.

### 2.2 Recruitment and screening

Participants with CAD will be recruited from two public hospitals in Granada, Spain: the “Hospital Universitario Virgen de las Nieves” and the “Hospital Universitario Clínico San Cecilio”. Recruitment started in May 2022, and we plan to enroll a minimum of 90 patients, aged 50-75, with stable CAD (phase III) proven by invasive coronary angiography or CT with at least one coronary stenosis with > 50% luminal reduction. A complete list of eligibility criteria is presented in **Table 1**. Eligible patients will be screened by the clinical and research team in three steps (**Figure 1**): 1) screening by the clinicians of the research team (i.e., based on clinical history); 2) screening based on a phone call conducted by the research team; and 3) screening during the baseline assessments including the cardiopulmonary exercise test (CPET) and echocardiography. Participants will be recruited in waves, with 10 to 20 people in each wave, throughout the year. In accordance with the CONSORT flow diagram (33), we will note the number of patients that were assessed for eligibility by the clinical and research team, the number of excluded patients (indicating reason for exclusion) during the enrollment phase and the number of randomized participants. For the allocation, the number of participants allocated to the intervention groups and the number of participants who received the intervention (with reasons for nonadherence to the allocated arm) will be noted. The number of participants who were lost to follow-up (dropouts) and who discontinued the intervention (including reasons) will be reported. Finally, the number of participants that were included in the analyses using the intention-to-treat and per-protocol databases will be described together with the reasons for the exclusion from the analytic database.

**Table 1.**
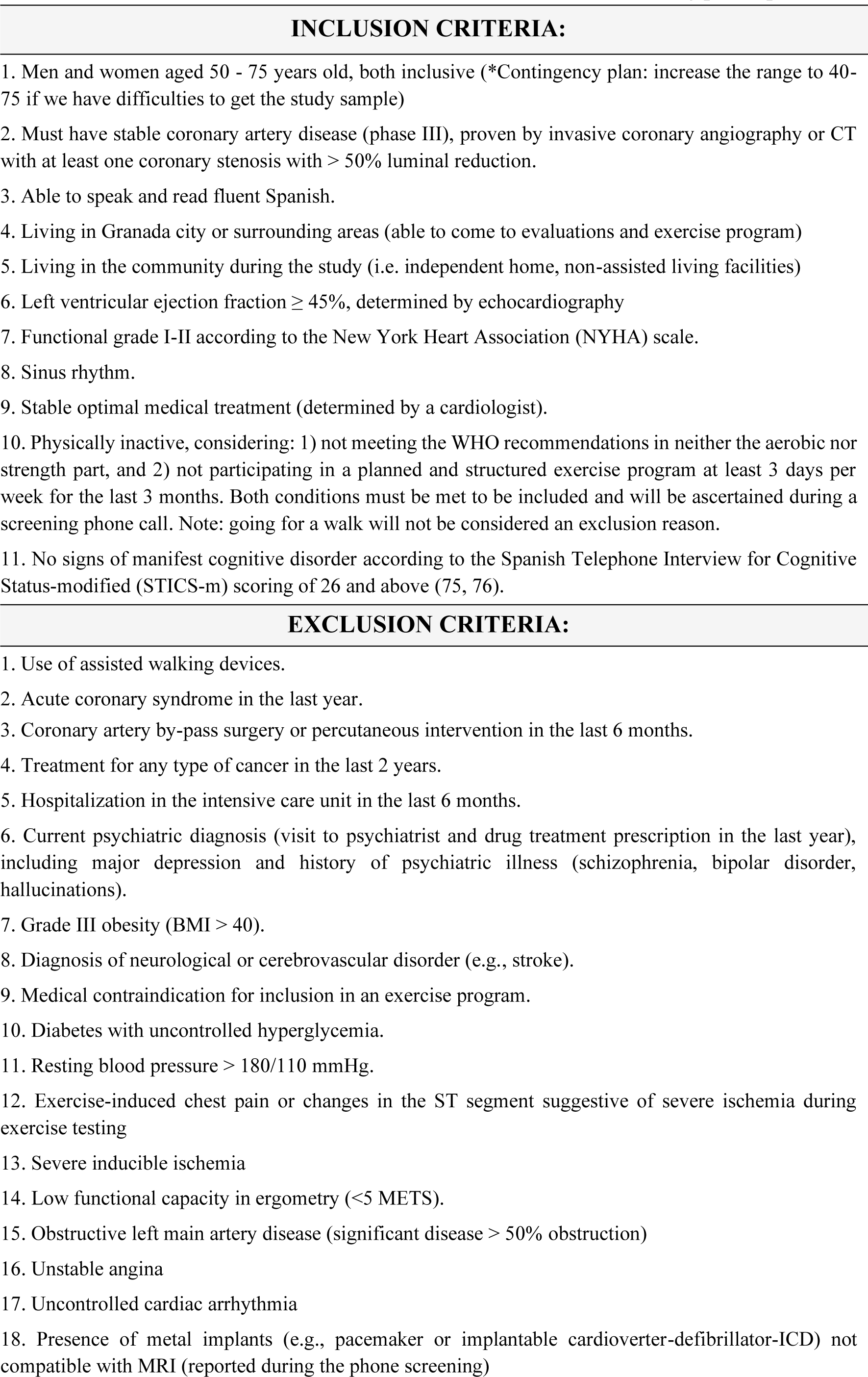
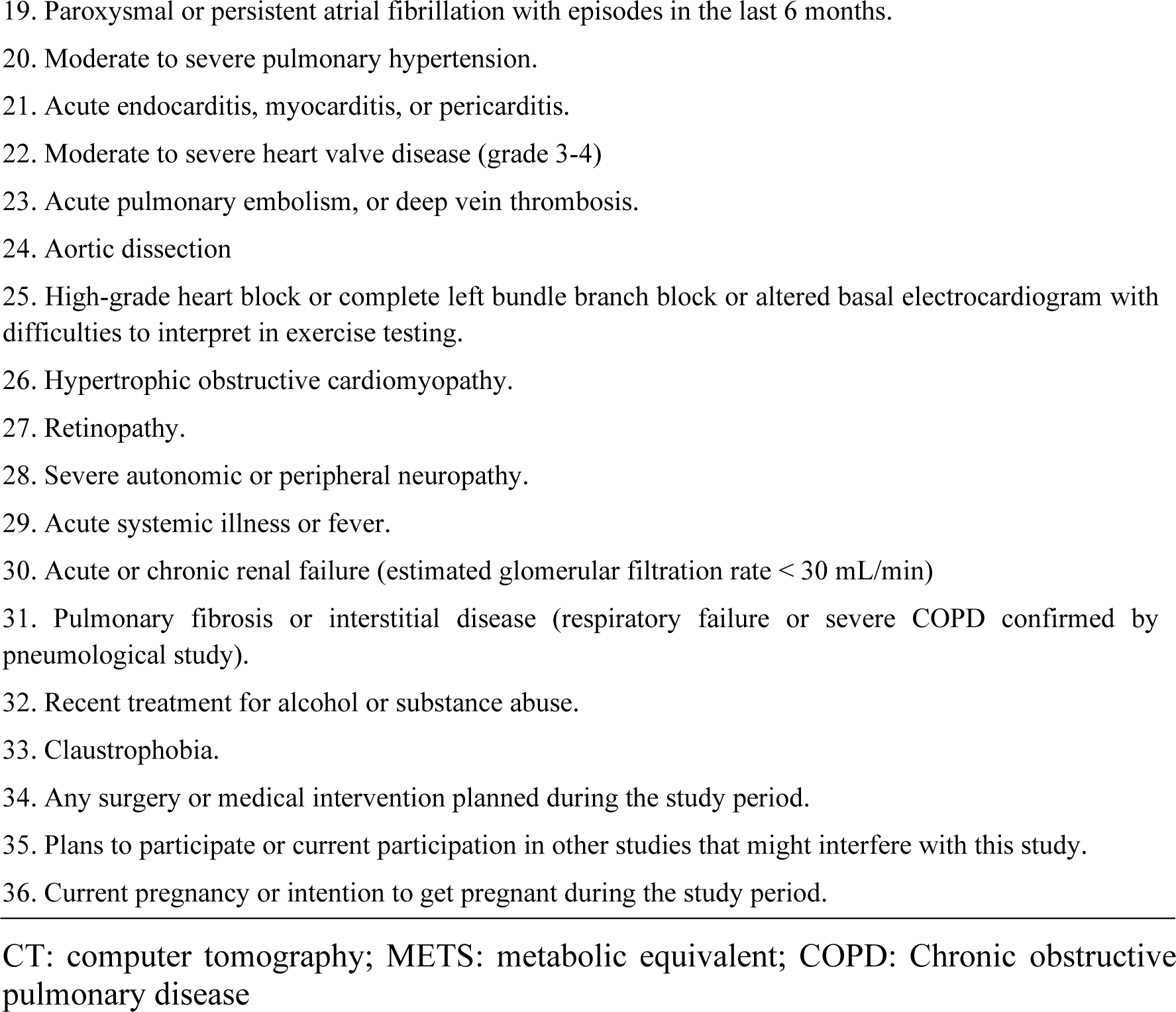
HEART-BRAIN inclusion and exclusion criteria for selecting participants.

### 2.3 Randomization

Randomization will occur on a rolling basis and only after the completion of all baseline assessments, through REDCap Software (34). Participants will be randomized using a 1:1:1 ratio and stratified by age (<65 years or ≥ 65 years) and sex (female or male). To ensure adequate allocation concealment, this procedure will be carried out by a blinded external researcher (Dr. Cabanas-Sanchez, Autonomous University of Madrid, Spain).

### 2.4 Blinding

Participants cannot be blinded due to the nature of the interventions, as they will inevitably become aware of their group allocation. Nonetheless, the principal investigator (PI) and the research team members participating in the post-intervention evaluations (who are not involved in monitoring the exercise sessions) will be blinded to the group assignments. Unblinding will only occur in case of an unexpected adverse event involving a participant and will only be done by the PI or a physician.

### 2.5 Interventions

A detailed and broader description of the program will be further extended in the Consensus on Exercise Reporting Template (CERT) study protocol of the HEART-BRAIN trial (*manuscript in preparation*). Briefly, participants will be randomized to one of the following three groups:

#### Aerobic high-intensity interval training (HIIT)

12-week duration, 3 times/week. This consists of a 4×4 HIIT (preferably on the treadmill): 4 intervals of 4 minutes at high intensity (85-95% of HRmax), interspersed with 3 intervals of 3 minutes of active resting at ∼70% of HRmax. All sessions include 10 minutes of warm-up and 10 minutes of cool-down, resulting in a total session time of 45 minutes. During the familiarization period (first two weeks), participants will progress from moderate-intensity training to the target HIIT.

#### Aerobic HIIT plus resistance training

12-week duration, 3 times/week. The aerobic part consists of a 3×4 HIIT (preferably on treadmill), with 3 intervals of 4 minutes at high intensity (85-95% of HRmax), interspersed with 2 intervals of 3 minutes of active resting (∼70% of HRmax). The resistance part consists of 2 rounds of an 8-exercise circuit (a combination of upper and lower body exercises using elastic bands and body weight) with a ratio of 20 seconds of effort followed by 40 seconds of resting. Sessions will have 5 minutes of warm-up on the treadmill and 5 minutes of cool-down walking around the gym, for a total session time of 45 minutes.

#### Control group – usual care

The control group will be treated as usual in outpatient Phase III, which in Spain includes periodic medical revisions where the clinician may provide patients with lifestyle and clinical advice (e.g., inform about exercise limitations, if any, and nutrition recommendations, stress management) and medication control. In addition, in the control group, we will apply the wait-list strategy, offering participants the HIIT supervised program described above, once data collection for pre- and post-intervention assessments is completed.

All intervention groups will receive the same usual care. The HIIT program and the HIIT part of the combined HIIT plus resistance training program have been designed based on the guidelines for the delivery and monitoring of HIIT in clinical populations (35) and meet the aerobic recommendations of the WHO guidelines, as it surpasses the minimum of 75 minutes of vigorous intensity or 150 minutes of moderate intensity per week, or a combination of them (23). The combined HIIT plus resistance training program meets the aerobic and resistance components of the WHO recommendations, as it has 3 sessions of resistance training per week while WHO guidelines recommend at least 2 sessions involving major muscle groups. Both exercise programs have been designed to provide an estimated isocaloric workload (same energy expenditure) in terms of intensity and duration.

Exercise programs will take place in the Sport and Health University Research Institute (iMUDS) at the University of Granada. Accredited expert trainers with a BSc degree in sports sciences will lead all supervised sessions with a ratio of 1:1 (one trainer, one participant). Exceptionally (e.g., scheduling conflicts), a ratio of 1:2 will be used.

Both training programs are individually tailored. Exercise prescription, monitoring, and decisions for progression will be based on the percentage of HRmax and the 10-point Borg Rating of Perceived Exertion scale (RPE) (36). HRmax will be determined during the CPET with electrocardiogram (ECG) and gas exchange analyzer performed at baseline (see section 2.3.2). Furthermore, symptoms and blood pressure will be monitored before, during, and after the exercise sessions. Participants taking beta-blocker medications often have a blunted HR response (35), which could also affect the HRmax obtained in the CEPT. Thus, while the HRmax will be used as a reference for exercise intensity, participants will be encouraged to learn and use RPE. This indicator will be used as a reference in case of discordance between HR and RPE targets. In addition, the trainers will use their judgment to adapt the intensity, particularly during the resistance training, because RPE might be less reliable in inexperienced participants (35).

### 2.6 Adherence, attendance, and compliance

Since the term adherence has been used with different meanings in the literature, in our study we will refer to two concepts more unequivocally used: attendance and compliance. Attendance will be defined as the percentage of sessions attended by the participant (recorded by the trainers) divided by the number of exercise sessions offered (12 weeks x 3 sessions/week = 36, yet holidays and logistic reasons may lead to slight deviations among waves and participants). Compliance will be defined as the percentage of sessions in which the specified amount of time (i.e., HIIT 6.5 min and HIIT plus resistance training 4.5 min) is spent within the target HR intensity, divided by the number of exercise sessions with valid data (e.g., sessions with technical issues of the HR bands will be excluded). The familiarization phase (i.e., the first two weeks of the exercise intervention) will not be considered for the compliance calculation. The amount of time, i.e., HIIT 6.5 min; HIIT plus resistance training 4.5 min, in the target HR intensity is based on the exercise protocol and the guidelines for HIIT prescription and monitoring in clinical populations (35): “for the first high-intensity interval, allow the entire 4-minute period to reach the HR target zone, for subsequent high intensity intervals (i.e., 2nd, 3rd, and/or 4th), allow 2-minutes (halfway) to reach the HR target zone”.

All protocol deviations will be recorded and reported (e.g., change in pre-defined inclusion/exclusion criteria, baseline and post assessments, data cleaning/processing).

### 2.7 Safety and adverse events

Although HIIT has been shown to be low risk in patients with CAD (37–39), the safety of our participants is the highest priority in the study. For these purposes, we followed the guidelines for using HIIT in clinical populations and have applied in our study the recommended exclusion criteria (35). Further, CPET together with cardiac and vascular ultrasound will be performed at baseline and serve as a clinical assessment to confirm the absence of underlying conditions that contraindicate exercise.

The center where the exercise training sessions will be delivered (iMUDS) is located in the Hospital area next to the emergency unit (200 meters) and the ambulance unit (600 meters). A trained allied health professional (nurse or physician) will be present in the center during the training sessions to guarantee a timely response in case of an adverse event. The center has a fully equipped trolley with a first aid kit and automatic defibrillator and obtained the Andalusian Certification of “Cardioprotected Center” (Decreto 22/2012 of February 14th). Furthermore, all members of the project team will receive certified training courses in cardiopulmonary resuscitation and the use of a semi-automatic defibrillator.

In case of an adverse event, we will follow the protocol recommended by the Spanish Council of Cardiopulmonary Resuscitation (CERCP). The number and reasons for adverse events (e.g., falls, injuries, musculoskeletal problems, major cardiovascular events, and any other events potentially related to the implementation of the trial protocol) occurring at any time during the study will be collected, reported, and separately described for each study arm. The participants allocated to the exercise group will be able to communicate any adverse events during their regular contact with the project team (i.e., 3 sessions/week), whereas those allocated to usual care will be asked to report any adverse event promptly, and will be called by phone for these regards at weeks 3, 9 and 12. Adverse events will be described in a table in the main HEART-BRAIN manuscript. Each adverse event will be clinically ascertained to judge whether the event was due to exercise in the corresponding study arm using an adverse event algorithm (i.e., Liverpool Causality Assessment Tool) (40) and recorded according to the Common Terminology Criteria for Adverse Events, (CTCAE) (41) by an experienced cardiologist (EME) after discussion and consensus with the rest of the cardiologists involved in the project.

### 2.8 Outcomes

The primary, secondary, and additional outcomes, the assessment instruments, and the data collection time points are summarized in **Table 2**. All outcome-related measures, data processing, and statistical analyses will be performed by staff blinded to the intervention assignment.

**Table 2.**
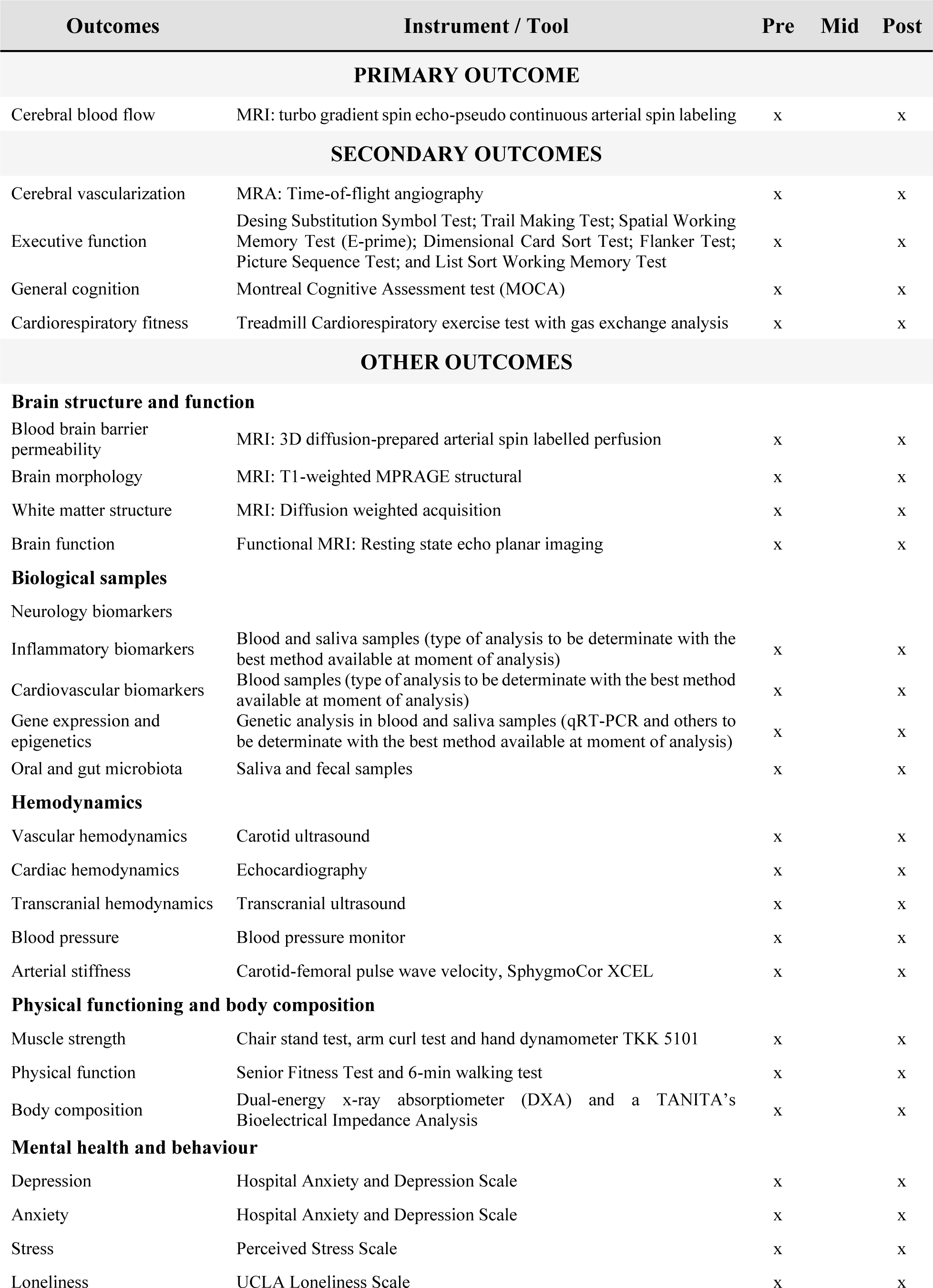
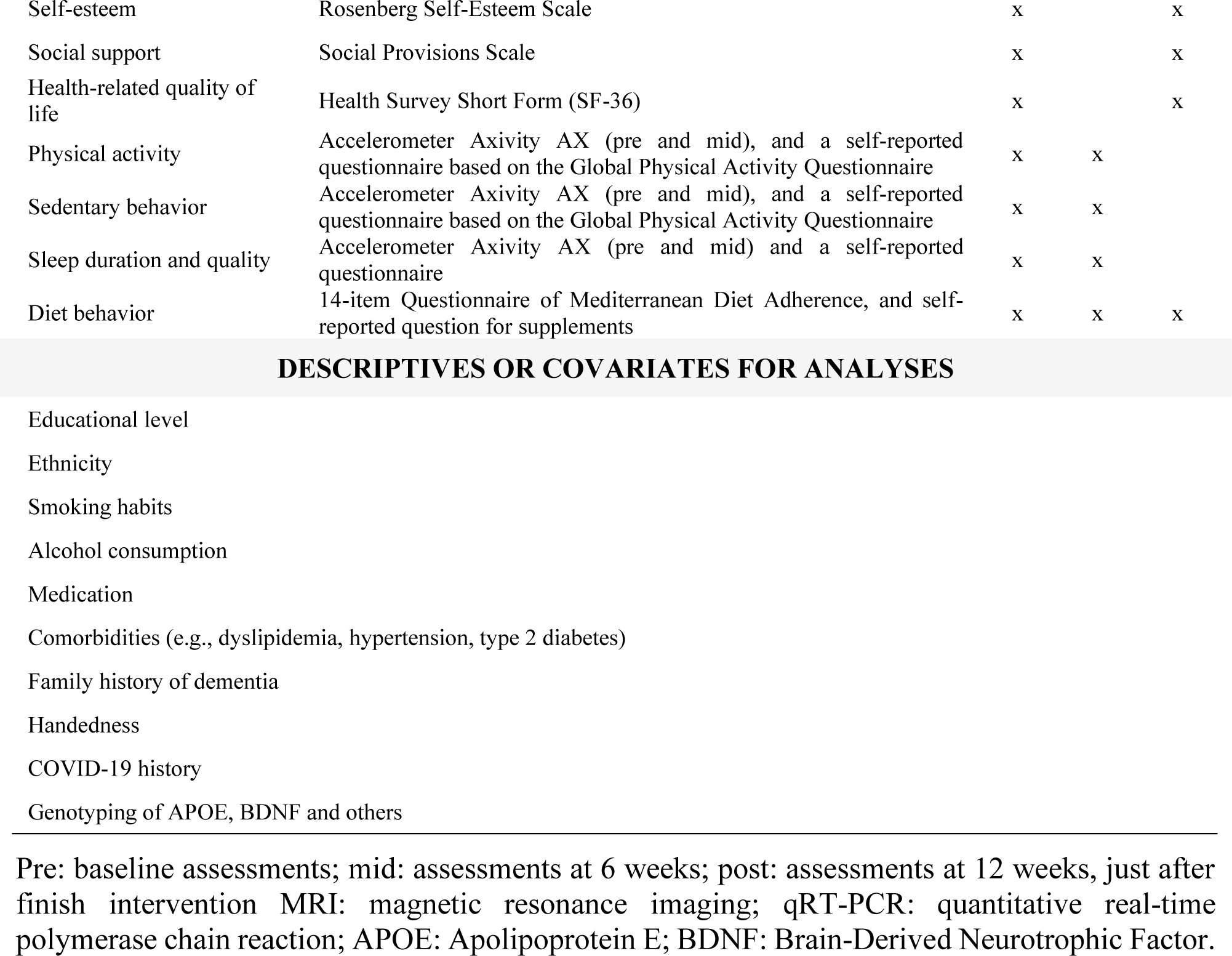
Outcomes instruments and time points of data collection.

#### 2.8.1. Primary Outcome Measures

##### Change in cerebral blood flow (timepoint: baseline and 12-weeks)

CBF (mL/100 g/min) will be measured using the MRI technique of pseudo-continuous arterial spin labeling (pCASL) (42, 43). We will analyze both global and regional CBF, as determined by voxel-wise analysis to measure local perfusion (acquisition parameters in **Table 3**).

**Table 3.** Brain Magnetic Resonance Imaging (MRI) parameters of the HEART-BRAIN trial.

| SEQUENCE | PARAMETERS | OUTCOMES | ACQUISITION TIME (MIN) |
| --- | --- | --- | --- |
| TOF (Time of flight angiography) | TR=21 ms, TE=3.43 ms, Flip angle: 18grad., FOV=200 mm, acquisition bandwidth of 155 Hz/PX, 44 slices (18.2% oversampling), resolution = 0.3×0.3×0.8 mm<sup>3</sup> | Angiography (cerebral vascularization) | 8 |
| T1-weighted MPRAGE structural | Sagittal, 0.8 mm isotropic resolution, TE/TI/TR = 2.31/1060/2400 ms, FOV = 256 mm, 224 slices | Brain structure | 6 |
| diffusion-prepared pCASL (DP-pCASL) sequence | TR = 4 sec, TE = 36.5 ms, FOV = 224 mm, matrix size = 64×64, 12 slices (10% oversampling), resolution = 3.5×3.5×8 mm<sup>3</sup>, label/control duration = 1500 ms, centric ordering, and optimized timing of background suppression for gray matter (GM) and white matter (WM) | Blood brain barrier permeability | 10 |
| Resting state EPI | Resolution: 2.5 × 2.5 × 2.5 mm, TE/TR = 40/1000 ms, Multiband factor = 8 (CMRR EPI sequence), 64 slices, 480 measurements | Resting BOLD (Blood Oxygen Level Dependent) | 5 |
| Diffusion weighted acquisition | Resolution: 2 × 2 × 2 mm, TE/TR = 95.6/2800 ms, Multiband factor = 4, b-values of 1500, 3000 s/mm<sup>2</sup>, 64 gradient directions | White matter microstructure | 8 |
| pCASL TGSE | 3D GRASE pCASL sequence, Resolution: 3.1 × 3.1 × 2.5 mm, TE/TR = 23.64/4300 ms, 48 slices, Multiple Post-labeling delay [0.5, 0.5, 1, 1, 1.5, 1.5, 2.0, 2.0, 2.0 s], Background Suppression, 9 measurements for labeling and control, 4 segment readout | Cerebral blood flow | 6 |
| MRI-Abdominal | Resolution: 2 × 2 × 2 mm, TE1-2/TR = 1.23-2.46/5.21 ms, flip angle 9°, CAIPIRINHA iPAT factor = 4, FOV = 448 mm | Visceral adipose tissue | 5 |

FOV: Field of View; Hz: hertz; PX: pixel; MPRAGE: Magnetization Prepared Rapid Gradient Echo; pCASL: pseudo-Continuous Arterial Spin Labelling; TE: Echo Time; TI: Inversion Time; TR: Repetition Time; EPI: Echo Planar Imaging; TGSE: Turbo Gradient Spin-Echo

#### 2.8.2 Secondary Outcome Measures

##### Change in cerebral vascularization (baseline to 12 weeks)

Cerebral vascularization will be measured using the time-of-flight-magnetic resonance angiography (TOF-MRA). This sequence allows imaging flow within vessels revealing the cerebrovascular anatomy without the need to administer contrast. Thus, the analysis of TOF-MRA provides the number, distribution, and morphology of intracranial vessels (acquisition parameters in **Table 3**).

##### Change in cognitive outcomes (baseline to 12 weeks)

A comprehensive neuropsychological battery will assess several domains of cognition (**Table 4**). General cognition will be assessed by the Montreal Cognitive Assessment (MoCA) test (44). Additionally, the battery includes paper-and-pencil tests (Trail making test (45) and Digit Symbol Substitution Test (46)), National Institutes of Health (NIH) Toolbox tests (Dimensional Change Card Sort Test, Flanker Test, Picture Sequence Memory Test, and List Sort Working Memory Test) (47) and a programmed test in E-prime software (Spatial Working Memory Test) (48). Detailed information about the cognitive tests has been previously described (49). The cognitive indicators will be transformed into standardized z-scores by calculating the difference between each individual raw score and the group average score at baseline, and then dividing by the standard deviation at baseline. We will determine the effects of the interventions on the cognitive outcomes (e.g., general cognition, cognitive flexibility, inhibition control, working memory, processing speed and episodic memory). To guarantee reliable evaluations, all paper-and-pencil tests will be independently scored by two trained evaluators, with any discrepancies resolved through mutual agreement.

**Table 4.** Cognitive tests included in the HEART-BRAIN trial.

| **Cognitive test** | **Day / session** | **Format** | **Domain** | **Time** |
| --- | --- | --- | --- | --- |
| Spanish Telephone Interview for Cognitive Status (75, 76) | Telephone screening | Verbal | General cognition | 10 min |
| Montreal Cognitive Assessment (44) | 2 | Verbal-paper-pencil | General cognition | 10 min |
| Trail Making Test A and B (45) | 2 | Paper-pencil | Executive function: Cognitive flexibility | 5 min |
| Digit Symbol Substitution Test (46) | 2 | Paper-pencil | Processing speed and attention | 5 min |
| Dimensional Change Card Sort Test (47) | 1 | Computerized | Executive function: Cognitive flexibility and inhibition | 5 min |
| List Sorting Working Memory Test (47) | 1 | Computerized | Executive function: Working memory (verbal) | 5 min |
| Flanker Test (47) | 1 | Computerized | Executive function: inhibitory control and attention | 5 min |
| Spatial Working Memory Test (48) | 3 | Computerized | Executive function: Working memory (spatial) | 10 min |
| Picture Sequence Memory Test (47) | 1 | Computerized | Episodic memory | 5 min |

##### Change in cardiorespiratory fitness (baseline to 12 weeks)

Cardiorespiratory fitness will be assessed by a standardized ramp CPET to exhaustion on a treadmill (h/p/cosmos, Nussdorf, Germany), in accordance with the American College of Sports Medicine Guidelines (50). We decided to use a ramp protocol with small increments, and opted for a standardized, instead of individualized, protocol to be able to compare the time-to-exhaustion (test duration, excluding warm-up) between participants as an additional indicator of exercise capacity and cardiorespiratory fitness, in addition to the peak oxygen uptake. CPET will be carried out by trained staff and under the supervision of a physician. The protocol begins with a 3-minute warm-up phase, during which the speed gradually increases to 4.8 km/h. Following this, the slope will be increased by 1% every 30 seconds until exhaustion or any other indication for test cessation (50), followed by a 3-minute recovery phase. This ramp protocol equates the 3-min increment Bruce tests, as indicated in the American College of Sports Medicine Guidelines (50). The respiratory exchange will be monitored with a dilution flow system (Omincal, Maastricht Instruments, Maastricht, the Netherlands), generating output data at 5-second intervals. A 12-lead electrocardiogram (AMEDTEC ECGpro, GmbH, Aue, Germany) will be continuously monitored by a physician during the test. Heart rate will also be registered with a Polar H10 monitor (including chest band and watch). Systolic and diastolic blood pressures will be measured with an automatic sphygmomanometer (Tango M2; SunTech Medical Inc., NC, USA) every 3 minutes during exercise, and after 3 and 5 minutes of recovery. The RPE will be asked every 3 minutes during the test (51).

Peak oxygen uptake (Peak V O_2_) will be defined as the highest rolling 30-second average during the test. Since the heart rate response might be blunted due to beta-blockers and the oxygen uptake plateau is not always observed, staff will note whether participants reach a respiratory exchange ratio (RER) of 1.1 or higher, as RER is one of the most robust indicators of maximal effort in these patients.

#### 2.8.3 Other Outcome Measures

##### Brain structure and function

CBF (primary outcome), cerebral vascularization (secondary outcome), along with other brain structure and function outcomes will be assessed by MRI at the Mind, Brain and Behavior Research Centre (CIMCYC) at the University of Granada. A Siemens Magnetom PRISMA Fit 3T scanner with a 64-channel head coil will be used. **Table 3** shows the imaging sequences that will be performed for each participant in acquisition order. Participants will be asked to wear comfortable clothing, MRI eligibility criteria will be checked (e.g., not having metallic implants or claustrophobia), and participants will provide additional informed consent specifically for MRI scans. When doubts exist about the compatibility of certain implants, an experienced radiologist will check the composition and material of the implant and inform about the compatibility with MRI before the scanning takes place. Standard MRI sequences will be conducted for approximately 60 minutes. A radiologist will review structural sequences to detect any incidental findings. If any of them are discovered, the radiologist will communicate with the participant for further examination. Besides CBF and cerebral vascularization, the following brain MRI outcomes will be examined (acquisition parameters are shown in **Table 3**):

##### Change in blood-brain barrier (BBB) permeability (baseline and 12 weeks)

BBB permeability will be assessed by a recently developed neuroimaging technique that measures water exchange across the BBB using 3D diffusion-prepared arterial spin-labelled perfusion MRI (52).

##### Change in brain morphology (baseline and 12 weeks)

MRI will measure brain morphology including volume, area, cortical thickness, and shapes by a T1-weighted MPRAGE (Magnetization Prepared Rapid Gradient Echo) structural sequence.

##### Change in white matter structure (baseline and 12 weeks)

MRI will measure white matter microstructure by a diffusion-weighted acquisition sequence.

##### Change in brain function (baseline and 12 weeks)

MRI will measure brain function during resting state and measures of brain activity and connectivity will be calculated.

#### Biological samples

The HEART-BRAIN trial aims to examine various biomarkers by the analysis of blood, saliva, and fecal samples. A trained nurse will collect blood samples from participants under fasting conditions (08:00 to 10:00 a.m.) at the Virgen de las Nieves University Hospital. For post-assessments, blood extraction will be done within the first 3-5 days after the last training session. Briefly, three ethylenediaminetetraacetic acid (EDTA) tubes, two citrate tubes, two serum tubes, and one PAXGENE tube will be collected corresponding to a total of approximately 30 ml of blood per participant visit. Part of the blood samples obtained from each participant will be processed directly at the Hospital (1 EDTA, 1 citrate, and 1 serum tube), and the remaining samples will be aliquoted and stored at −80°C for later studies. Saliva samples will be collected into sterilized plastic containers and aliquoted into 1.5mL Eppendorf. Fecal samples will also be collected under standardized conditions using sterile plastic containers and stored at −80°C. **Table 5** outlines the specifications of the samples, initial analyses, and preliminary analysis plans. Nevertheless, samples will be analyzed using the best available methods at the time of processing. Blood analyses will cover conventional biochemical measurements, cardiovascular, brain-peripheral, and inflammatory biomarkers, telomere length, and genetic analysis. Saliva and fecal samples will undergo metagenomic analysis to study the microbiome. Some of the analyses will be conducted as additional funding is obtained. Briefly, the main outcomes planned from biological samples include:

**Table 5.** Biological samples and targeted analysis plan.

| SAMPLE | ANALYSIS TARGET | PRELIMINARY ANALYSIS PLAN |
| --- | --- | --- |
| **Blood samples** |  |  |
| Plasma | Aβ (Amyloid beta) peptides Aβ1-42, Aβ1-40; BD-tau and P-tau; Neurofilament light chain (NFL); Glial fibrillary acidic protein (GFAP); Vascular endothelial growth factor (VEGF); chemokine C-X-C motif ligand 13 (CXCL-13); IgM-1; IL-17; pancreatic polypeptide (PPY); Adiponectin; Brain-derived neurotrophic factor (BDNF); Cathepsin B; vascular cell adhesion molecule 1 (VACM-1); C-reactive protein (CRP); high-sensitivity CRP; Interleukin-6 (IL-6); Interleukin-17 (IL-17); Tumor necrosis factor (TNF-Alpha); Klotho; GLP-1, Clusterin | Plasma biomarker Ab (Amyloid beta) peptides Ab1-42, Ab1-40; BD-tau and P-tau; Neurofilament light chain - NFL GFAP- concentrations will be measured using Single molecule array (Simoa) methods on an HD-X instrument and commercial assays from Quanterix. Key inflammatory biomarkers will be analyzed with automated blood cell counters and quantified by multiple analyte profiling technology (MILLIPLEX R MAP Human High Sensitivity T-Cell Magnetic Bead Panel, EMD Millipore Corporation, Missouri, U.S.A.) Klotho will be determined using a solid-phase sandwich enzyme-linked immunosorbent assay (Demeditec, Kiel, Germany) according to the manufacturer's protocol. |
| Peripheral Blood Mononuclear Cells | Telomere length | A commercial DNA isolation kit (Qiagen, Barcelona, Spain) will be used to extract genomic DNA from isolated PBMCs. Relative telomere length will be determined by quantitative real-time polymerase chain reaction (qRT-PCR) using the telomere/single-copy gene ratio (T/S) |
| Serum | Insulin-like growth factor 1 (IGF-1); Insulin; Glucose; Glycated Hemoglobin (HbA1c), creatinine, Serum fatty acid (SFA-1), total, HDL and LDL cholesterol, glomerular filtration rate, triglycerides | IGF-1, insulin, glucose, glycated hemoglobin (HbA1c) and serum fatty acids (SFA-1) will be analyzed using XMap technology (Luminex Corporation, Austin, TX) and human monoclonal antibodies (Milliplex Map Kit; Millipore, Billerica, MA). Total cholesterol, HDL-C, LDL-C, triglycerides, and glucose will be assessed using a spectrophotometer. Insulin will be assessed by chemiluminescence immunoassay. |
| Whole Blood | Glycated hemoglobin (A1c), RNA and DNA analyses | Blood specimens will be used for genotyping. The genotypes will be determined using TaqMan genotyping assays (e.g., APOE). A commercial RNA isolation kit (Qiagen, Barcelona, Spain) will be used to extract total RNA from whole blood samples. The selection of specific miRNAs will be based on the existing bibliographic information. In accordance with this, the selected miRNA will be evaluated using Taqman probes. |
| **Saliva samples** | Oral microbiota and other potential biomarkers | To be determined with the best method available at the moment of analysis |
| **Fecal samples** | Metagenomic Analysis of the Gut Microbiome | To be determined with the best method available at the moment of analysis |

##### Change in cardiovascular biomarkers (baseline and 12 weeks)

Blood samples will be used to determine concentrations of cardiovascular biomarkers including glucose, insulin, triglycerides, high-density lipoprotein (HDL), low-density lipoprotein (LDL), and total cholesterol.

##### Change in brain-peripheral biomarkers (baseline and 12 weeks)

Blood and saliva samples will be used to determine the concentration of peripheral brain biomarkers such as brain-derived neurotrophic factor (BDNF), vascular endothelial growth factor (VEGF), insulin-like growth factor 1 (IGF-1), cathepsin B (CTSB), amyloid and tau protein levels, as well as novel neurodegenerative biomarkers based on new evidence available at the time of the analysis.

##### Change in inflammatory biomarkers (baseline and 12 weeks)

Blood samples will be used to determine concentrations of inflammatory peripheral biomarkers such as tumor necrosis factor-alpha (TNF-alpha), interleukins, and both traditional and high-sensitivity C-reactive proteins.

##### Change in gene expression and epigenetics (baseline and 12 weeks)

Blood samples will be stored for epigenetic and gene expression changes.

##### Change in oral and gut microbiota (baseline and 12 weeks)

Saliva and fecal samples will be used to determine oral and gut microbiota including the most representative phyla (i.e., firmicutes, Bacteroidetes, and proteobacteria).

#### Hemodynamics

##### Hemodynamic changes (baseline and 12 weeks)

Non-invasive hemodynamic parameters will be measured using ultrasound (Vivid E95, GE Healthcare) at three levels: 1) vascular (main carotid artery and vertebral artery, with measurements of intima-media thickness, flow, and wall strain, as well as registration of presence of atherosclerotic plaques), 2) cardiac (i.e., cardiac dimensions and volumes, systolic and diastolic function such as ejection fraction, strain deformation, and myocardial work), and 3) transcranial (i.e., systolic and diastolic flow velocity in the middle cerebral artery). Image acquisition and analysis will be carried out by cardiologists with expertise in ultrasonography.

##### Change in blood pressure (baseline and 12 weeks)

Resting systolic and diastolic blood pressure will be assessed by a validated automated blood pressure monitor (Omron M3, Intellisense, OMRON Healthcare Europe, Spain) in the sitting position after 5 minutes of rest and with the left arm resting on a table at heart level. Three readings will be collected with 2-minute intervals in between. Blood pressure will be calculated as the average of the last two blood pressure readings (53).

##### Change in arterial stiffness (baseline and 12 weeks)

Arterial stiffness will be assessed using the pulse wave analysis and carotid-femoral pulse wave velocity (aortic-PWV) by applanation tonometry determined by the SphygmoCor® XCEL PWA/PWV (AtCor Medical, Sydney, Australia).

##### Physical functioning and body composition

Additional physical function and body composition outcomes will include:

###### Change in physical function and fitness (baseline and 12 weeks)

The Senior Fitness Test, including the 6-min walking test) will be used to assess overall physical functioning. Senior Fitness Test battery assesses upper and lower body strength, aerobic capacity, walking speed, balance, and flexibility (54). Perceived physical fitness (cardiorespiratory fitness, muscular strength, speed-agility, flexibility and overall fitness) will be assessed using the International Fitness Scale (IFIS) (55). In addition, muscular strength with be measured with a combination of some of the Senior Fitness Test and handgrip strength test. In brief, each participant will be encouraged to perform the maximal isometric force twice with each hand using a hand dynamometer (TKK 5101 Grip D, Takey, Tokyo Japan). The maximum value of each hand will be taken and averaged in kilograms (kg) as an indicator of upper-body muscular strength. Lower body muscular strength will be assessed using the chair stand test (number of repetitions). Upper body muscular strength will be assessed using the arm curl test (number of repetitions). Perceived strength will be assessed using a single item from the IFIS (55).

###### Change in body composition (Baseline and 12 weeks)

Body mass index (BMI, kg/m^^2^) will be computed from height and weight measured with SECA instruments following standardized procedures. Body composition (i.e., lean mass, kg, fat mass, kg, and bone mineral content and density, z-score) will be assessed using a dual-energy x-ray absorptiometer and a TANITA’s Bioelectrical Impedance Analysis (TANITA MC-980MA-N plus, Amsterdam, the Netherlands). We will also conduct MRI of the abdominal region to obtain information on overall visceral adipose tissue, specific-organ adipose tissue, and subcutaneous abdominal adiposity.

#### Mental health and lifestyle behaviors

A battery of questionnaires will be administered to evaluate the following dimensions in relation to mental health and lifestyle behavior:

<u>Change in depressive symptoms</u> (baseline and 12 weeks) will be assessed using the Global Deterioration Scale (56), and the Hospital Anxiety and Depression Scale (HADS) (57); <u>change in anxiety symptoms</u> (baseline and 12 weeks) will be assessed using the HADS questionnaire (57); <u>change in stress</u> (baseline and 12 weeks) will be assessed using the Perceived Stress Scale (58); <u>change in loneliness</u> (baseline and 12 weeks) will be assessed using the UCLA Loneliness Scale (59); <u>change in self-esteem</u> (baseline and 12 weeks)will be assessed using the Rosenberg Self-Esteem Scale (60); <u>change in social support</u> (baseline and 12 weeks) will be assessed using the Social Provisions Scale (61). <u>Change in health-related quality of life</u> (baseline and 12 weeks) will be assessed using the SF-36 form (62), which provides an 8-scale profile of functional health and well-being scores (i.e., physical functioning; role limitations due to physical problems; bodily pain; general health perceptions; energy/vitality; social functioning; role limitations due to emotional problems; and mental health) as well as psychometrically-based Physical Component Score (PCS) and Mental Component Score (MCS).

<u>Change in physical activity and sedentary behaviors</u> (baseline, 6 and 12 weeks). Physical activity and sedentary behavior will be measured using the accelerometer Axivity AX at baseline and 6-weeks. The accelerometer will be attached to the non-dominant wrist for 9 consecutive days, the sampling frequency will be set at 100 Hz, and the accelerometer raw data processing will be conducted using open-source software tools to ensure transparency and replicability of the methods (63). Additionally, participants will complete the short version self-reported International Physical Activity Questionnaire(64) at baseline and 12 weeks.

<u>Change in duration and sleep quality</u> (baseline and 6 weeks). Sleep duration (hours/night) will be computed from the accelerometer Axivity AX and diaries, and an open-source software will be used to process the raw data recordings (63). In addition, participants will complete a single-item question on usual sleep duration. Sleep quality (e.g. sleep regularity, sleep efficiency, latency, number and total time of waking up after sleep onset) will also be assessed using the accelerometer Axivity AX.

<u>Change in diet behaviors</u> (baseline, 6 and 12 weeks). Diet behaviors will be self-reported using the 14-item Questionnaire of Mediterranean Diet Adherence (PREDIMED-14) with an additional question for supplement intake.

<u>Additional variables related to health, lifestyle and genotype</u> will be measured for descriptive purposes, for covariate adjustment in the analyses when necessary or for interaction with the intervention effects. Examples include biological sex, ethnicity, educational level, smoking habits, alcohol consumption, medication, comorbidities (e.g., dyslipidemia, hypertension, or type 2 diabetes), family history of dementia, handedness, and COVID-19 history. Additionally, genotyping at baseline will be conducted for well-known Single Nucleotide Polymorphisms (SNP) for brain health, such as Apolipoprotein E (APOE) and BDNF genotypes, and additional relevant SNPs based on an updated literature search. The genotyping analysis is conducted to test gene-exercise interaction as we have done in previous RCTs (49).

### 2.9 Sample size and statistical analysis

A detailed description of the statistical analysis plan, including a sample size calculation, for the HEART-BRAIN trial has been pre-registered and published on Clinicaltrials.gov (NCT06214624; submission date: January, 2024).

#### 2.9.1 Sample size

The HEART-BRAIN project has been designed to detect a low-to-medium size change (i.e. effect size Cohen d=0.35) with an alpha of 0.05 and beta of 0.2 (power 0.8). The trial has been powered for a 7% dropout during the intervention (based on our previous RCTs (49)). Thus, the estimated sample size, for repeated-measures ANOVA using G*Power (version 3.1.9.7), was 90 participants (30 in each study arm).

#### 2.9.2 Framework and timing

A superiority hypothesis testing framework will be used. In the main analyses, we will compare whether both exercise interventions (HIIT only or HIIT plus resistance training) are superior to usual care (no exercise training). The final analyses will be performed after collecting and processing the data of the primary and secondary outcomes. The time points for data collection of each outcome are explained in section 2.4.

#### 2.9.3 Statistical interim analyses and stopping guidance

No pre-specified interim analyses will be performed, and therefore, a stopping guidance is not applicable. Basic analyses (not including the primary outcome) have been performed as required by the final report for the funding agencies, after completion of the first 58 participants. The basic analyses have been performed by a researcher who is not involved in data collection (EAB). The results of the basic analyses have not been discussed with other members of the research team involved in the evaluations or interventions to avoid any bias during the implementation of the study, data collection or processing. These basic analyses have not led to any protocol deviations.

#### 2.9.4 Brief description of the primary and secondary analyses

The main analyses will consist of the intention-to-treat analyses for the primary and secondary outcomes using a constrained baseline (meaning baseline adjusted) linear mixed model, which accounts for baseline differences among the study groups. The model will include fixed effects for time (two levels) and treatment (three levels) as well as the unique participant identifier as a random effect. If the global test of significance indicates between-group differences, pairwise comparisons will be explored. Although no adjustments for multiplicity will be performed, family-wise type 1 error rate on the primary outcome will be retained by using a hierarchical analytic approach. The prespecified hierarchical hypotheses will be tested using the prespecified sequence: HIIT plus resistance training versus usual care, HIIT versus usual care, and HIIT plus resistance training versus HIIT. No additional adjustments for covariates will be made (except for baseline adjustments of the outcome). The main statistical analyses will be performed by an independent researcher (EAB), who is not involved in the recruitment, evaluations, and interventions, and who will be blinded to treatment allocation by coding the intervention arms (e.g., A, B, C). The same approach will be applied to secondary and tertiary outcomes.

#### 2.9.5 Moderation, mediation, and other secondary analyses

Although the RCT is not powered for subgroup analyses, we will perform moderation analyses by exploratory subgroup analyses for age, sex, education level, baseline level of the study outcome (stratified on the median), and APOE genotype. Likewise, we plan to explore potential mechanisms driving effects on outcomes. For example, we will run a formal mediation analysis to test whether changes in cardiorespiratory fitness explain exercise-derived changes in brain and cognitive outcomes, and whether changes in CBF or vascularization mediate exercise-derived changes in cognitive outcomes.

#### 2.9.6 Confidence intervals and P values

All statistical tests will be two-tailed. P for significance will be set at 0.05 and 95% confidence intervals will be estimated. No adjustments for multiplicity will be made as multiplicity adjustments may be of lesser importance in the case of distinct treatment arms (65). Since only one primary outcome has been defined, other outcomes do not require adjustment for multiple testing. However, we will include a hierarchical analytic approach for the exercise interventions tested and the primary outcome.

#### 2.9.7 Analysis populations

We plan to use two approaches for the statistical analyses:

1. Intention-to-treat (main analysis): This will be used for our primary analyses and includes all randomized participants. With this approach, all randomized participants are included in the analysis, based on the groups to which they were initially randomly assigned to. This implies that some participants will have valid data at both time points, while some might have missing data at baseline or post-intervention assessment, which is sometimes named as “available-case intention-to-treat”.
2. Per-protocol and additional sensitivity analyses: This dataset will be used for our secondary analyses and includes all participants with ≥ 70% attendance. Additionally, sensitivity analyses will be performed using the per-protocol database and excluding participants who had injuries or other health problems during the post-assessment evaluations. We will also analyze the effects of attendance and compliance by conducting an analysis including individuals who, in addition to attending ≥70% of the sessions, also had ≥70% compliance to the training protocol based on the time within the HIIT heart rate target zones.

### 2.10 Data management and sharing

The HEART-BRAIN trial will use two data storage systems: (i) the REDCap platform, an online software for the management of research databases in clinical trials and translational research. RedCap will be used for the storage and management of all non-imaging information, guarantying its trace and safety (34); (ii) the HEART-BRAIN desktop computer will also contain participants’ pseudonymized data collected on paper-based forms, that will be scanned and stored, as well as the MRI data. Physical copies of data collection forms and documents will be securely stored in a locked cabinet at the iMUDS facility. Upon enrolment in the trial, participants will be assigned a unique code to be used consistently across all forms and data collection reports, i.e. pseudonymized data, to reduce the risk of exposure resulting from unintended unauthorized access or disclosure. In addition, information linking the pseudonym to the identifiable information must be kept separately and subjected to ensure non-attribution to an identified or identifiable individual. The PI will have access to the final dataset and will provide reasonable and responsible access to the pseudonymized data to other researchers. Participants will consent to the use of their pseudonymized data for secondary research, as outlined in the informed consent process. Once a participant is enrolled, all data will undergo thorough quality control checks and be archived following the storage protocol at iMUDS. Quality control measures include visual inspection of imaging data, double correction of paper-pencil cognitive tests, and verification of data validity and integrity in REDCap. The study will adhere to the FAIR principles (Findability, Accessibility, Interoperability, and Reusability), and meta-data will be made publicly available. A DOI (Digital Object Identifier) is issued to every published record. Governance, ethical considerations, and shared trial oversight will be prioritized, aligning with current best practices. Study results, including positive, negative, or inconclusive, will be disseminated through peer-reviewed journals, and national and international conferences, and shared via social media and press releases with participants, caregivers, physicians, and the broader medical and scientific community.

The protocol, statistical analyses plan and data management plan will be shared open access. Data files, including pseudonymized individual participant data, will be shared under restricted access and upon reasonable request (contact FBO) due to privacy issues and EU-GDPR regulations. In principle, all collected individual participant data will be available for sharing under the “as open as possible, as closed as necessary” principle. The shared data files will be pseudonymized, and only include participants who provided informed consent for sharing, and sharing is only possible when the data is used for research purposes related to exercise and cardiovascular and brain health. The individual participant data will be available 12 months after the primary outcome paper is published. The specific process of data access will be determined in a later stage. In short, data will be made available upon reasonable requests to the PI (FBO). The data requests must contain the purpose and aim of the research, target variables, and data analysis plan before data sharing. Data will only be shared for research and public health purposes. A data access committee will be created to approve the appropriate data requests. A data monitoring committee does not exist in the funding system in Spain.

## 3 DISCUSSION

CAD is associated with cognitive and mental health impairment (7–10). However, the mechanisms explaining the connection between the heart and the brain in patients with CAD remain unknown (9). Growing evidence supports the benefits of exercise on the cardiovascular health of individuals with CAD (66–71). Yet, there is scarce knowledge on its influence on brain health. The mechanisms by which different types of exercise might improve brain health or attenuate the cognitive and mental health declines observed in patients with CAD remain to be elucidated. Well-designed intervention studies unraveling the effects and mechanisms by which exercise impacts brain health in patients with CAD will have important clinical and scientific implications. The findings will facilitate moving towards a more ‘personalized medicine’ approach to exercise prescription in patients with CAD. Thus, the present work provides a comprehensive description of the HEART-BRAIN RCT, following the SPIRIT guidelines for interventional RCTs (30–32). The HEART-BRAIN project aims to examine the effects of two different 12-week supervised exercise interventions (HIIT and HIIT combined with resistance training) compared to usual care on brain and cardiovascular health in patients with CAD. The primary outcome is CBF assessed by a cutting-edge technique called ASL-MRI.

Among the few studies that have evaluated the effects of exercise on brain health in individuals with CAD, most of them have focused on health-related quality of life and mental health outcomes (72, 73). Other key domains of brain health such as cognition, biological markers of brain health, brain morphology and function, or cerebrovascular outcomes remain unexplored.

This study will contribute to the advancement of scientific knowledge in different ways. Firstly, it stands as an innovative intervention of two different exercise programs in accordance with the most updated guidelines (23, 35, 50, 51). By providing a comprehensive description and pre-specified plan of the study protocol, we ensure transparency and enable the scientific community to replicate the protocols and intervention, fostering the dissemination of scientific knowledge. We expect that the intervention and the obtained results will serve as a valuable resource for healthcare and exercise professionals, allowing them to effectively prescribe individualized exercise to patients with CAD. This represents an effective transfer of scientific knowledge to practical settings. This study underscores the importance of systematic and well-defined methodological reports addressing the transparency gaps within the scientific community and facilitating the comprehensive sharing of study information.

### 3.1 Strengths and limitations

HEART-BRAIN has several strengths that bolster the reliability and validity of the expected results, such as: i) well-designed RCT framework ensuring random allocation of participants, minimizing selection bias, and enhancing the validity of the causal inferences drawn from the data; ii) extensive brain health mapping, including comprehensive assessments of the different domains of brain health; iii) comprehensive set of variables, including objective, gold-standard test (e.g., CPET) and patient-reported outcomes (e.g., QoL) to encompass both physical and brain health; iv) continuous heart rate monitoring throughout the exercise sessions allowing for a rigorous compliance analysis, v) time and (estimated) caloric matched exercise groups, ensuring that any observed effects can be attributable to the type of exercise rather than differences in exercise duration or intensity; vi) flexibility is offered to help participants overcome barriers frequently reported about exercise participation (e.g., trainers adapt to the time that fits best participants’ schedule); vii) our mechanistic approach will provide insights on how exercise can impact brain health in patients with CAD and the potential mechanisms explaining the heart-brain connection, such as changes in CBF; and viii) this study holds significant clinical relevance as it specifically targets potential improvements in brain health among patients with CAD, a population known to experience accelerated cognitive decline.

The study also has some limitations that must be acknowledged. Since the HEART-BRAIN RCT is implementing HIIT, certain criteria have been established to maximize safety. For example, only individuals with a stable and controlled disease during the outpatient phase will be included. Future trials recruiting a more generalizable group of patients with CAD (i.e., including individuals in a more severe condition) are needed. Further, our study is of relative short duration (12 weeks). While notable physiological changes may manifest within this timeframe, certain significant adaptations may require a longer intervention and follow-up period. Although inherent to exercise-based RCTs, another limitation is the single (and not double) blinding. These limitations should be taken into consideration when interpreting the study’s findings.

## Supporting information

Supplemental Table 1. SPIRIT checklist

## Data Availability

All data produced in the present study are available upon reasonable request to the authors

## 4 Acknowledgments

We thank Dr. Veronica Cabanas-Sanchez for performing the randomization of the study.

## 6 Author Contributions

Conceptualization and project direction: FBO (principal investigator); Project coordination: AT; Writing - original draft: AT, FBO; Writing - review & editing: AT, PSU, EAB, IMF, SVA, AC, IEC, FBO. Formal analysis: EAB, PSU, FBO. All authors contributed intellectually to the design and development of the methodology and approved the final manuscript of this study protocol. Our authorship aligns with the ICMJE author contributions criteria (74).

