## Supplemental Table 1. SPIRIT checklist for "Exercise and brain health in patients with coronary artery disease: study protocol for the HEART-BRAIN randomized controlled trial"

### Supplementary table S1. SPIRIT Checklist of the HEART-BRAIN Trial.

SPIRIT — STANDARD PROTOCOL ITEMS: RECOMMENDATIONS FOR INTERVENTIONAL TRIALS

SPIRIT-Outcomes 2022 Checklist
(for combined completion of SPIRIT 2013 and SPIRIT Outcomes 2022 items)

| Section | Item No | SPIRIT 2013 and 2022 items | Addressed headings; pags. |
|---|---|---|---|
| **Administrative information** | | | |
| Title | 1 | Descriptive title identifying the study design, population, interventions, and, if applicable, trial acronym | Title page; 1 |
| Trial registration | 2a | Trial identifier and registry name. If not yet registered, name of intended registry | Trial registration; 1<br>Design and ethics; 7 |
| | 2b | All items from the World Health Organization Trial Registration Data Set | N/A |
| Protocol version | 3 | Date and version identifier | Title page; 1<br>Design and ethics; 7 |
| Funding | 4 | Sources and types of financial, material, and other support | Funding; 30 |
| Roles and responsibilities | 5a | Names, affiliations, and roles of protocol contributors | Title page; 1<br>Author contributions; 29<br>Funding; 30 |
| | 5b | Name and contact information for the trial sponsor | Trial sponsor; 1 |
| | 5c | Role of study sponsor and funders, if any, in study design; collection, management, analysis, and interpretation of data; writing of the report; and the decision to submit the report for publication, including whether they will have ultimate authority over any of these activities | Conflict of interest; 29<br>Funding; 30 |

| | | | |
|---|---|---|---|
| | 5d | Composition, roles, and responsibilities of the coordinating centre, steering committee, endpoint adjudication committee, data management team, and other individuals or groups overseeing the trial, if applicable (see Item 21a for data monitoring committee) | N/A |
| **Introduction** | | | |
| Background and rationale | 6a | Description of research question and justification for undertaking the trial, including summary of relevant studies (published and unpublished) examining benefits and harms for each intervention | Introduction; 4-6 |
| | 6b | Explanation for choice of comparators | Introduction; 4-6 |
| Objectives | 7 | Specific objectives or hypotheses | Study objectives; 6 |
| Trial design | 8 | Description of trial design including type of trial (eg, parallel group, crossover, factorial, single group), allocation ratio, and framework (eg, superiority, equivalence, noninferiority, exploratory) | Design and ethics; 7<br>Study objectives; 6<br>Randomization; 8-9<br>Framework and timing; 23 |
| **Methods: Participants, interventions, and outcomes** | | | |
| Study setting | 9 | Description of study settings (eg, community clinic, academic hospital) and list of countries where data will be collected. Reference to where list of study sites can be obtained | Design and ethics; 7<br>Recruitment and screening; 8 |
| Eligibility criteria | 10 | Inclusion and exclusion criteria for participants. If applicable, eligibility criteria for study centers and individuals who will perform the interventions (eg, surgeons, psychotherapists) | Recruitment and screening; 8<br>Table 1; 38-39 |
| Interventions | 11a | Interventions for each group with sufficient detail to allow replication, including how and when they will be administered | Interventions; 9-11 |
| | 11b | Criteria for discontinuing or modifying allocated interventions for a given trial participant (eg, drug dose change in response to harms, participant request, or improving/worsening disease) | Safety and adverse events; 12-13 |

| | | | |
|---|---|---|---|
| | 11c | Strategies to improve adherence to intervention protocols, and any procedures for monitoring adherence (eg, drug tablet return, laboratory tests) | Interventions; 9-11<br>Adherence, attendance, and compliance; 11-12 |
| | 11d | Relevant concomitant care and interventions that are permitted or prohibited during the trial | Interventions; 9-11 |
| Outcomes | 12 | Primary, secondary, and other outcomes, including the specific measurement variable (eg, systolic blood pressure), analysis metric (eg, change from baseline, final value, time to event), method of aggregation (eg, median, proportion), and time point for each outcome. Explanation of the clinical relevance of chosen efficacy and harm outcomes is strongly recommended | Outcomes; 13-22<br>Table 1; 38-39<br>Framework and timing; 23 |
| | 12.1 | Provide a rationale for the selection of the domain for the trials primary outcome | Introduction; 4-6 |
| | 12.2 | If the analysis metric for the primary outcome represents within-participant change, define and justify the minimal important change in individuals | Introduction; 4-6<br>Outcomes; 13-22 |
| | 12.3 | If the outcome data collected are continuous but will be analyzed as categorical (method of aggregation), specify the cutoff values to be used | Outcomes; 13-22 |
| | 12.4 | If outcome assessments will be performed at several time points after randomization, state the time points that will be used for analysis | Outcomes; 13-22<br>Table 2; 40-41 |
| | 12.5 | If a composite outcome is used, define all individual components of the composite outcome | Outcomes; 13-22 |
| Participant timeline | 13 | Time schedule of enrolment, interventions (including any run-ins and washouts), assessments, and visits for participants. A schematic diagram is highly recommended (see Figure) | Recruitment and screening; 8<br>Figure 1; 37 |
| Sample size | 14 | Estimated number of participants needed to achieve study objectives and how it was determined, including clinical and statistical assumptions supporting any sample size calculations | Sample size and statistical analysis; 22-27 |
| | 14.1 | Define and justify the target difference between treatment groups (eg, the minimal important difference) | Confidence intervals and P values: 24 |
| Recruitment | 15 | Strategies for achieving adequate participant enrolment to reach target sample size | Recruitment and screening; 8 |

**Methods: Assignment of interventions (for controlled trials)**

Allocation:

| | | | |
|---|---|---|---|
| Sequence generation | 16a | Method of generating the allocation sequence (eg, computer-generated random numbers), and list of any factors for stratification. To reduce predictability of a random sequence, details of any planned restriction (eg, blocking) should be provided in a separate document that is unavailable to those who enrol participants or assign interventions | Randomization; 8-9 |
| Allocation concealment mechanism | 16b | Mechanism of implementing the allocation sequence (eg, central telephone; sequentially numbered, opaque, sealed envelopes), describing any steps to conceal the sequence until interventions are assigned | Randomization; 8-9 |
| Implementation | 16c | Who will generate the allocation sequence, who will enrol participants, and who will assign participants to interventions | Randomization; 8-9 |
| Blinding (masking) | 17a | Who will be blinded after assignment to interventions (eg, trial participants, care providers, outcome assessors, data analysts), and how | Blinding; 9 |
| | 17b | If blinded, circumstances under which unblinding is permissible, and procedure for revealing a participant's allocated intervention during the trial | Blinding; 9<br>Safety and adverse events; 12-13<br>Brief description of the primary and secondary analyses: 23-24 |

**Methods: Data collection, management, and analysis**

| | | | |
|---|---|---|---|
| Data collection methods | 18a | Plans for assessment and collection of outcome, baseline, and other trial data, including any related processes to promote data quality (eg, duplicate measurements, training of assessors) and a description of study instruments (eg, questionnaires, laboratory tests) along with their reliability and validity, if known. Reference to where data collection forms can be found, if not in the protocol | Outcomes; 13-22<br>Table 2; 40-41<br>Data management and sharing; 25-27 |
| | 18a.1 | Describe what is known about the responsiveness of the study instruments in a population similar to the study sample | Introduction; 4-6<br>Sample size and statistical analysis; 22-27 |
| | 18a.2 | Describe who will assess the outcome (eg, nurse, parent) | Outcomes; 13-22 |

| | | | |
|---|---|---|---|
| | 18b | Plans to promote participant retention and complete follow-up, including list of any outcome data to be collected for participants who discontinue or deviate from intervention protocols | Adherence, attendance, and compliance; 11-12 Analysis populations; 25 |
| Data management | 19 | Plans for data entry, coding, security, and storage, including any related processes to promote data quality (eg, double data entry; range checks for data values). Reference to where details of data management procedures can be found, if not in the protocol | Data management and sharing; 25-27 |
| Statistical methods | 20a | Statistical methods for analysing primary and secondary outcomes. Reference to where other details of the statistical analysis plan can be found, if not in the protocol | Sample size and statistical analysis; 22-27 |
| | 20a.1 | Describe any planned methods to account for multiplicity in the analysis or interpretation of the primary and secondary outcomes (eg, coprimary outcomes, same outcome assessed at multiple time points, or subgroup analyses of an outcome) | Confidence and P values; 23 |
| | 20b | Methods for any additional analyses (eg, subgroup and adjusted analyses) | Sample size and statistical analysis; 22-27 |
| | 20c | Definition of analysis population relating to protocol non-adherence (eg, as randomised analysis), and any statistical methods to handle missing data (eg, multiple imputation) | Sample size and statistical analysis; 22-27 |
| **Methods: Monitoring** | | | |
| Data monitoring | 21a | Composition of data monitoring committee (DMC); summary of its role and reporting structure; statement of whether it is independent from the sponsor and competing interests; and reference to where further details about its charter can be found, if not in the protocol. Alternatively, an explanation of why a DMC is not needed | Data management and sharing; 25-27 |
| | 21b | Description of any interim analyses and stopping guidelines, including who will have access to these interim results and make the final decision to terminate the trial | Sample size and statistical analysis; 22-27 |
| Harms | 22 | Plans for collecting, assessing, reporting, and managing solicited and spontaneously reported adverse events and other unintended effects of trial interventions or trial conduct | Safety and adverse events; 12-13 |
| Auditing | 23 | Frequency and procedures for auditing trial conduct, if any, and whether the process will be independent from investigators and the sponsor | N/A |
| **Ethics and dissemination** | | | |
| Research ethics approval | 24 | Plans for seeking research ethics committee/institutional review board (REC/IRB) approval | Design and ethics; 7 |

| | | | |
|---|---|---|---|
| Protocol amendments | 25 | Plans for communicating important protocol modifications (eg, changes to eligibility criteria, outcomes, analyses) to relevant parties (eg, investigators, REC/IRBs, trial participants, trial registries, journals, regulators) | Design and ethics; 7 |
| Consent or assent | 26a | Who will obtain informed consent or assent from potential trial participants or authorised surrogates, and how (see Item 32) | Design and ethics; 7 |
| | 26b | Additional consent provisions for collection and use of participant data and biological specimens in ancillary studies, if applicable | Design and ethics; 7<br>Biological samples; 17-18 |
| Confidentiality | 27 | How personal information about potential and enrolled participants will be collected, shared, and maintained in order to protect confidentiality before, during, and after the trial | Data management and sharing; 25-27 |
| Declaration of interests | 28 | Financial and other competing interests for principal investigators for the overall trial and each study site | Funding; 30 |
| Access to data | 29 | Statement of who will have access to the final trial dataset, and disclosure of contractual agreements that limit such access for investigators | Data management and sharing; 25-27 |
| Ancillary and post-trial care | 30 | Provisions, if any, for ancillary and post-trial care, and for compensation to those who suffer harm from trial participation | Design and ethics; 7<br>Interventions; 9-11 |
| Dissemination policy | 31a | Plans for investigators and sponsor to communicate trial results to participants, healthcare professionals, the public, and other relevant groups (eg, via publication, reporting in results databases, or other data sharing arrangements), including any publication restrictions | Data management and sharing; 25-27 |
| | 31b | Authorship eligibility guidelines and any intended use of professional writers | Data management and sharing; 25-27 |
| | 31c | Plans, if any, for granting public access to the full protocol, participant-level dataset, and statistical code | Data management and sharing; 25-27 |
| **Appendices** | | | |
| Informed consent materials | 32 | Model consent form and other related documentation given to participants and authorised surrogates | Design and ethics; 7 |

| | | | |
|---|---|---|---|
| Biological specimens | 33 | Plans for collection, laboratory evaluation, and storage of biological specimens for genetic or molecular analysis in the current trial and for future use in ancillary studies, if applicable | Design and ethics; 7<br>Biological samples; 17-18<br>Table 5; 44 |

*It is strongly recommended that this checklist be read in conjunction with the SPIRIT 2013 Explanation & Elaboration for important clarification on the items. Amendments to the protocol should be tracked and dated. The SPIRIT checklist is copyrighted by the SPIRIT Group under the Creative Commons "Attribution-NonCommercial-NoDerivs 3.0 Unported" license.
